# Post Traumatic Stress Disorder in Patients with Traumatic Spinal Cord Injury

**DOI:** 10.1101/2024.08.11.24311824

**Authors:** Mahdi Haq, Nabhan Rashad

**Affiliations:** NeuroCareAI; Khyber Teaching Hospital

## Abstract

**Introduction:** Post traumatic stress disorder (PTSD) is a psychiatric disorder that may develop after exposure to a traumatic event. Patients of traumatic spinal cord injuries are at risk of developing PTSD, and diagnosing this disorder and recognizing risk factors is important for effective treatment.

**Objective:** To determine the prevalence of PTSD in post-traumatic spinal cord injury patients and correlate the presence of PTSD to factors such as age, cause of injury, and level of injury.

**Methods:** A descriptive cross sectional study was conducted at Paraplegic Center in Peshawar, Pakistan. The Diagnostic and Statistical Manual of Mental Disorders Fifth Edition (DSM-5) was used to assess the presence of post-traumatic stress disorder in patients at the Paraplegic Center. The study was carried out from 20 December 2014 to 20 February 2015 on a convenience sample of 51 patients. The criterion for inclusion in the study was to have a traumatic spinal cord injury, while the exclusion criterion was to have a spinal cord injury that was non-traumatic in nature.

**Results:** Out of 51 patients, 31% met the diagnostic criteria for PTSD. The age group of 15-24 years had a 27% prevalence of PTSD, while the age groups of 25-34 years and 35-44 years had a PTSD prevalence of 42% and 40% respectively. Patients who had fallen from a height had the highest prevalence of PTSD – 41%, as compared to patients who had other causes of traumatic spinal cord injury. Patients with a lumbar spinal lesion had a PTSD prevalence of 44%, whereas patients with a cervical and thoracic spinal lesion had a PTSD prevalence of 33% and 25% respectively.

**Conclusion:** The study shows that the middle age groups had a higher prevalence of PTSD, and patients who had fallen from a height had the highest prevalence of PTSD. Lumbar spinal lesion patients had a higher prevalence of PTSD than patients who had spinal lesions at the cervical or thoracic level.

## BACKGROUND

### Introduction

PTSD is a psychiatric disorder that may develop after exposure to a traumatic event, or a series of traumatic events. These traumatic events may include exposure to events such as warfare, serious injuries, sexual assault, or other events which pose a threat to life. ^[1]^

According to the United States Department of Veteran Affairs,

“PTSD can occur after you have been through a traumatic event. A traumatic event is something terrible and scary that you see, hear about, or that happens to you, like:

● Combat exposure
● Child sexual or physical abuse
● Terrorist attack
● Sexual or physical assault
● Serious accidents, like a car wreck
● Natural disasters, like a fire, tornado, hurricane, flood, or earthquake”.

Furthermore, Mayo Clinic defines this disorder as the following:

“PTSD is a mental health condition that’s triggered by a terrifying event — either experiencing it or witnessing it.”

Traumatic events and deaths of dear ones are common in peoples’ lives. In a study conducted by WHO covering 21 different countries, greater than 10% of the people surveyed reported experiencing a traumatic event. 21.8% reported that they witnessed violence, while 18.8% said that they experienced interpersonal violence, 17.7% were involved in accidents, 16.2% were exposed to war, and 12.5% witnessed trauma to a loved one. It is estimated that 3.6% of the entire world’s population has suffered from Post-traumatic Stress Disorder at some point in their life. ^[2]^

PTSD occurs regardless of age, whether young or old. It has been shown that women are at a greater risk of developing post-traumatic stress disorder as compared to men. There is some evidence that the susceptibility of a person to have post-traumatic stress disorder may be increased if the disorder runs in the family.

PTSD afflicts people regardless of their age. It commonly occurs in war veterans, and victims of physical and sexual assault, mental or physical abuse, accidents, and natural or man-made disasters. Having said this, it is not necessary that a person with PTSD has gone through a dangerous event. PTSD may occur in a person even if someone they know, such as a friend or a relative, goes through a dangerous experience. Furthermore, PTSD can be caused by the sudden and unexpected death of a loved one. ^[3]^

PTSD can present with a wide array of symptoms, which can be categorized into three main groups as follow:

**“**1. Re-experiencing symptoms

● Flashbacks—reliving the trauma over and over, including physical symptoms like a racing heart or sweating
● Bad dreams
● Frightening thoughts.

2. Avoidance symptoms

● Staying away from places, events, or objects that are reminders of the experience
● Feeling emotionally numb
● Feeling strong guilt, depression, or worry
● Losing interest in activities that were enjoyable in the past
● Having trouble remembering the dangerous event.

3. Hyperarousal symptoms

● Being easily startled
● Feeling tense or “on edge”
● Having difficulty sleeping, and/or having angry outbursts. Children and teens can have extreme reactions to trauma, but their symptoms may not be the same as adults. In very young children, these symptoms can include:
● Bedwetting, when they’d learned how to use the toilet before
● Forgetting how or being unable to talk
● Acting out the scary event during playtime
● Being unusually clingy with a parent or other adult.” ^[4]^

Going through a traumatic event, especially one that results in paraplegia or quadriplegia can be a tough ordeal. People may undergo a range of reactions and emotions following trauma, with most people recovering given adequate time. Having said that, a small number of people develop severe, chronic psychological issues after the trauma. This study will help us understand the emotional disturbances that are felt by people following a traumatic event, and furthermore will let us see what percentage of people progress into having PTSD. There is a stigma associated with mental health, and this study will help raise awareness about mental disorders such as PTSD. Research has shown that a combination of medication and therapy can be effective in treating PTSD. ^[5]^

### Literature Review

A study was conducted by M.S. Nielsen in Denmark from March to July 2001 in rehabilitation centers for spinal cord lesion patients. The sample size was 69 and the tool used for assessment of PTSD was the Harvard trauma questionnaire. The results depicted that PTSD was prevalent in 20% of the subjects. ^[6]^

In another study conducted by R. Bryant, J. Marosszeky, J. Crooks, and J. Gurka, and published by The American Journal of Psychiatry in April 2000, a sample size of 96 patients in a brain injury rehabilitation unit in the United States was interviewed based on the DSM-III-R criteria. PTSD was diagnosed in 27.1% patients with traumatic brain injury. ^[7]^

A study was conducted at a spinal injuries center in Stoke Mandeville Hospital in the United Kingdom by P. Kennedy and M.J. Evans. The total sample size was 85 with an average age of 32.6. Out of the 85 subjects, 17 were female. Out of the total sample, 70% had complete lesions and 40% had tetraplegia. Road traffic accidents were the most common cause of injury followed by accidents related to sports. Level of distress was high in 14% of the patients. Trauma related distress was significantly higher in female patients or patients with a high level of anxiety or depression. ^[8]^

### Rationale of the Study

This study assessed the prevalence of PTSD in patients with traumatic spinal cord injuries, in hopes of recognizing the psychological effects of these injuries on the patients in order to raise awareness about providing proper psychological and/or psychiatric therapeutic services in a timely manner.

### Objectives

1. To determine the prevalence of PTSD in post-traumatic spinal cord injury patients.
2. To determine the frequency of PTSD in different age groups of post-traumatic spinal cord injury patients.
3. To determine the percentage of PTSD according to different causes of injury and different levels of spinal cord lesions in post-traumatic spinal cord injury patients.

## METHODOLOGY

### Location

The study was conducted at Paraplegic Centre in Peshawar, Pakistan.

### Study population

The patients admitted at the Paraplegic Centre with a spinal cord traumatic injury.

### Study design

It was a descriptive cross-sectional study.

### Study duration

Data was collected from 20^th^ December 2014 to 20^th^ February 2015.

### Sampling

Sampling size was 51 inpatients of traumatic spinal cord injury. Sampling scheme was convenience sampling.

### Data collection tool

The data collection tool that was used for detecting PTSD in the patient population was the PTSD diagnostic criteria from the The Diagnostic and Statistical Manual of Mental Disorders 5^th^ edition published by the American Psychiatric Association.

### Operational definitions

*Inclusion criteria:* Required the patient to be a traumatic spinal cord injury patient admitted in the Paraplegic Centre in Hayatabad, with at least a gap of one month between the occurrence of the injury and the interview of the patient.

*Exclusion criteria:* Outpatients who were there for follow-up, and patients who did not have traumatic spinal cord injury.

### Ethical considerations

Permission was taken from the administration of the Paraplegic Centre prior to starting the study. The participating students filled out the appropriate paperwork for permission to interview admitted patients at the Paraplegic Centre, and permission was duly granted.

Informed consent was taken prior to interviewing each patient, and measures were taken to ensure the patient’s privacy for the duration of the interview. The patients’ names were not published in the study, and were discarded after analysis of the data. Only the supervisor, investigator and co-investigators of the study had access to the filled questionnaires.

### Analysis plan

The data was analysed using Microsoft Excel.

## RESULTS

### Sample description

The study was conducted over a course of 2 months, from 20th December 2014 to 20th February 2015 at Paraplegic Center in Hayatabad Peshawar. A total of 51 patients were interviewed for the study during this time, of which 31% met the diagnostic criteria for PTSD. The range of ages for the positive PTSD patients interviewed was 18-46 years, and the average age was 28.8 years.

### Analysis

According to the study 16 out of 51 were positive for PTSD according to the DSM-5 scale. This comes out to be 31%.

**Figure 1:**
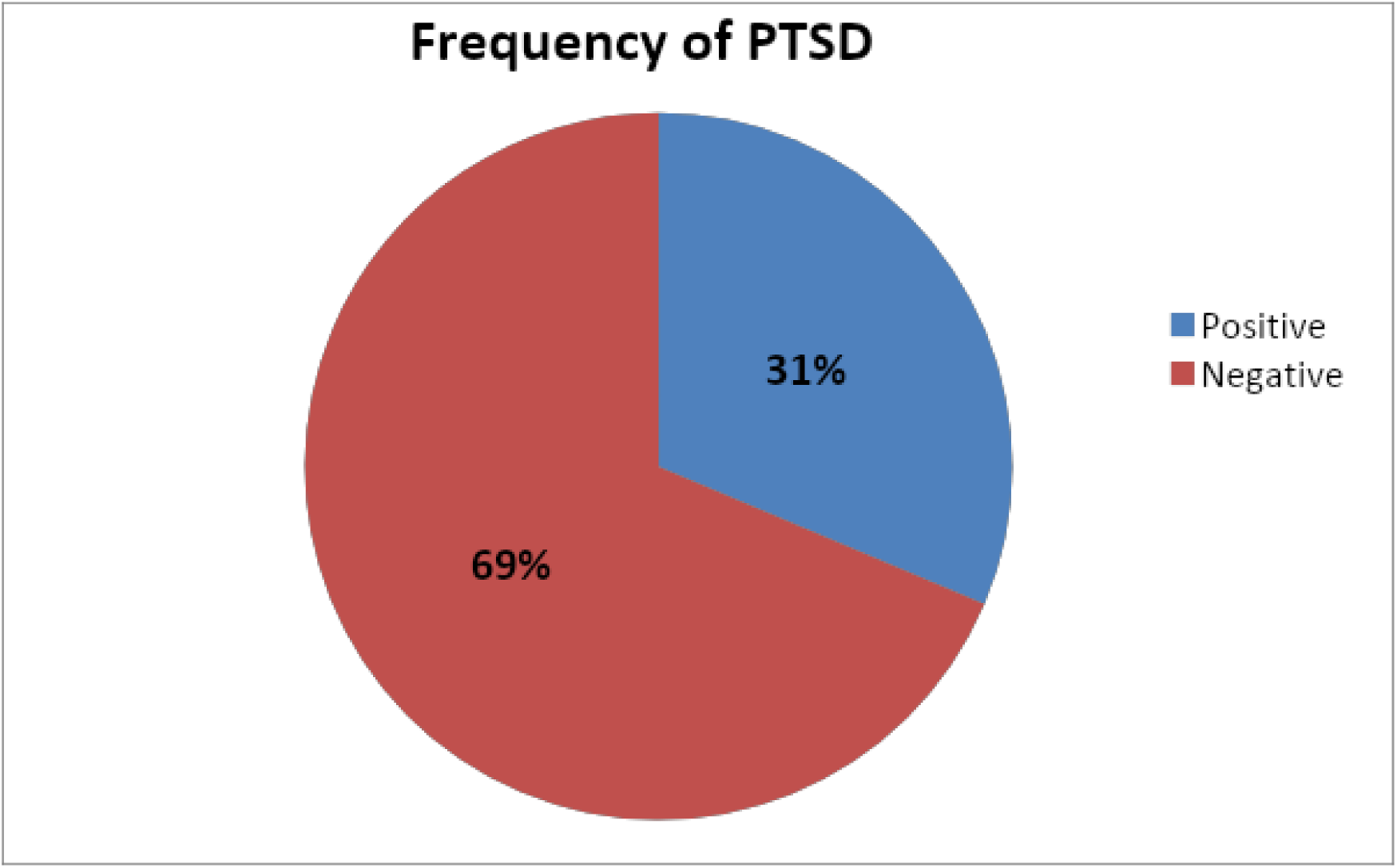
Overall frequency of PTSD in patients with traumatic spinal cord injury

**Figure 2:**
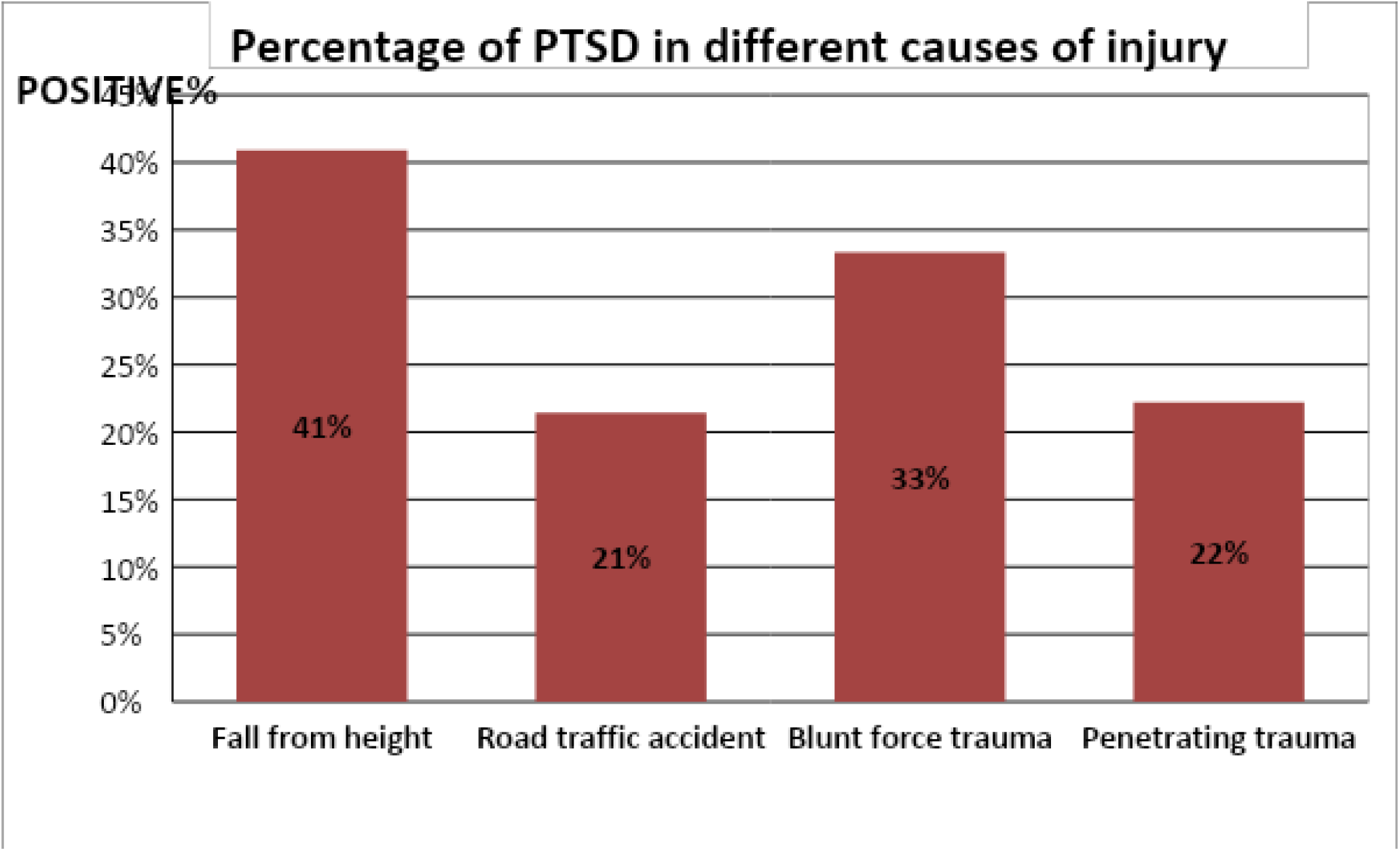
Percentage of PTSD in different cause of injury

The cause of injury was also found out to have a strong association with PTSD. PTSD was highest in subjects who experienced fall from height, (9 out of 22 [41%] had a history of fall from height) while 2 out of 6 (33%) of PTSD subjects had blunt force trauma. 22% of PTSD patients had a penetrating trauma while only 3 out of 14 (21%) with a history of road traffic accident were PTSD positive.

**Figure 3:**
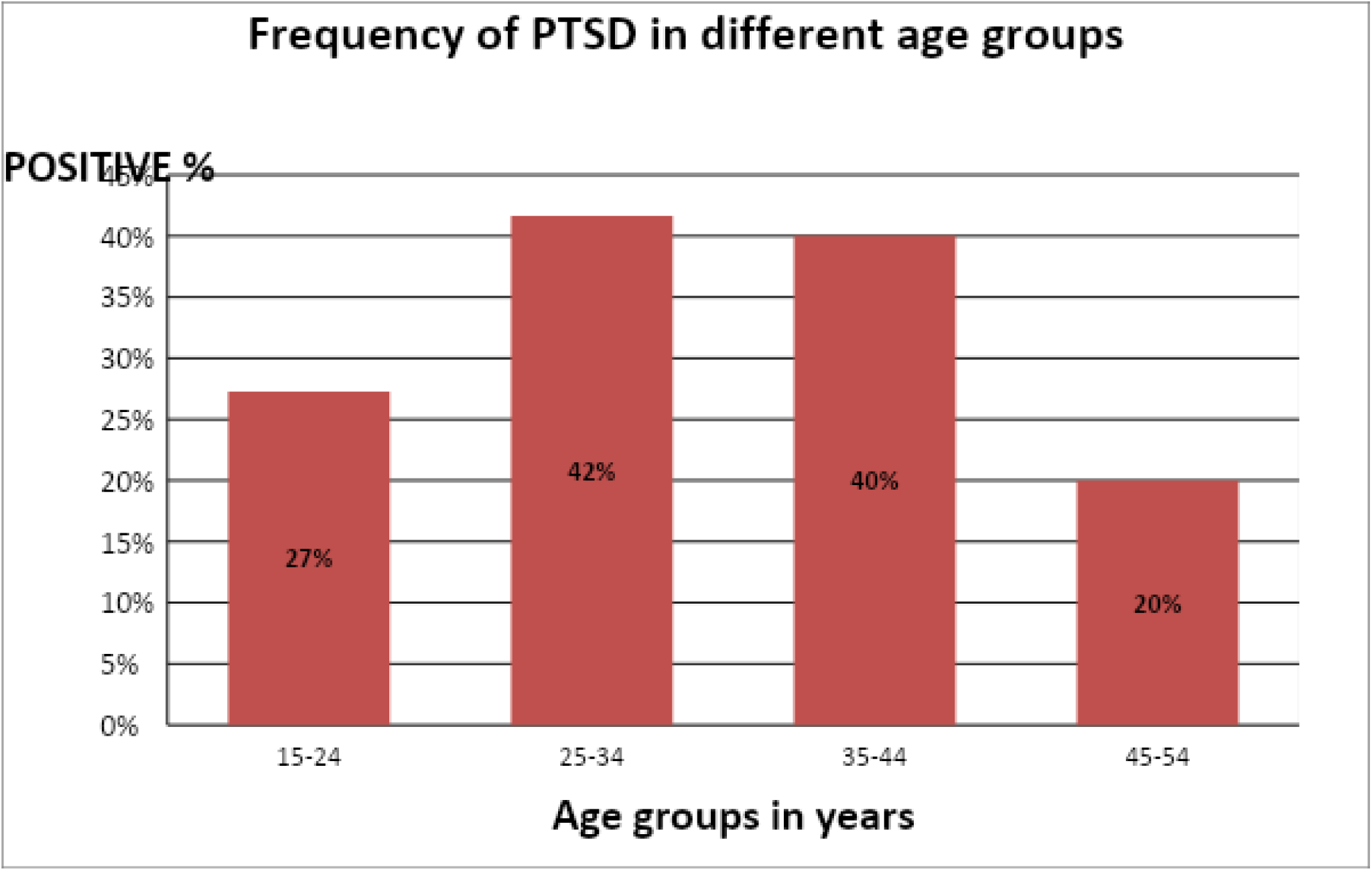
Frequency of PTSD in different age groups

The age of the subjects also had a strong association to PTSD. High percentage (42%) of PTSD was found in subjects of age group 25-34 years while less percentage (20%) was identified in 45 to 54 years of age group. In the age group 35-44 and 15-24, the PTSD was found to be 40% and 27% respectively.

**Figure 4:**
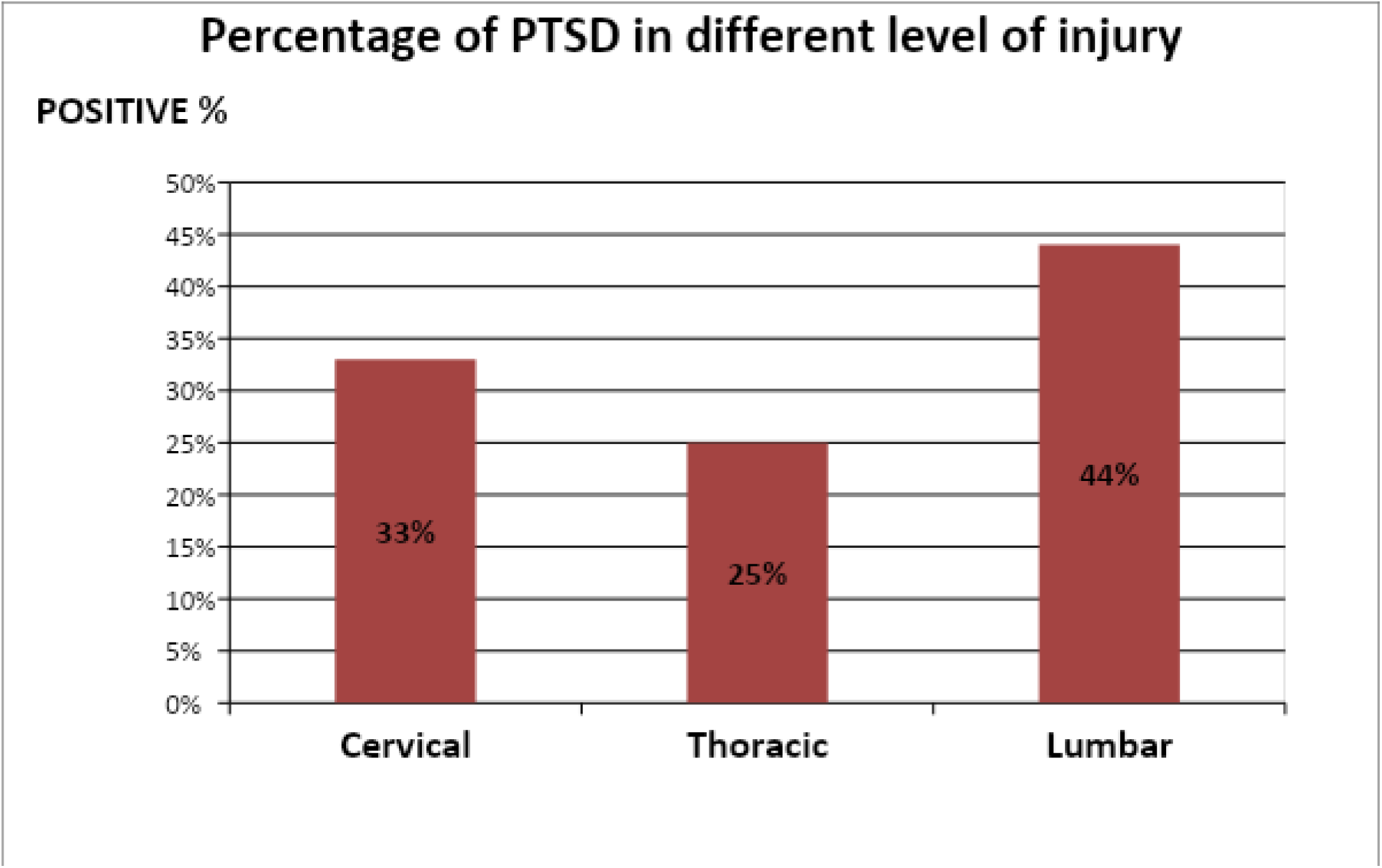
Percentage of PTSD in different level of injury

The prevalence of PTSD was assessed in relation to the level of injury, and it was found that 4 out of 9 subjects (44%) with spinal cord lesions at lumbar region were positive for PTSD while 4 out of 12 (33%) and 7 out of 28 (25%) with spinal cord lesions at cervical and thoracic regions respectively were found to be positive.

## DISCUSSION

The main aim of this study was to assess the status and frequency of PTSD in patients who suffered from trauma to the spinal cord in the past. They have been through a life threatening accident and are under a lot of stress. Trauma to spinal cord is generally associated with lifelong morbidity and disability that’s why they are more prone to develop disorders like depression and PTSD. A study conducted by MS Nielsen in Denmark March to July 2001 in rehabilitation centers for spinal cord lesion. Sample size was 69 and tool used for assessment of PTSD was Harvard trauma questionnaire. The results depicted that PTSD was prevalent in 20% of the subjects. ^[4]^ While our study showed that PTSD was prevalent in 31% of patients, there is a difference between the results which may be due to many reasons. The scale used is different in both of the studies, as we used DSM-5 criteria while Harvard trauma questionnaire was used in aforementioned study. The studies are conducted at entirely different setups, our study is conducted in Peshawar, Pakistan while the aforementioned study was conducted in Denmark which is a developed country with more facilities for such traumatic and handicapped patients e.g. psychiatric services which leave them less susceptible for disorders like PTSD, while our setup lags behind in providing such care to patients having lifelong traumatic lesions.

### Strengths

DSM V scale was used to assess status of the patients. DSM V is a widely used tool which is accepted globally and serves as a universal authority for psychiatric diagnosis.

### Limitations

The main limitation of the study is the convenience sampling method, which increases the probability of systemic sampling error, and reduces the power of the study. The presence of more than one researcher in the data collection procedure could affect the results and the consistency of the data collection tool. Caution should have been exercised to avoid drawing inferences.

The DSM V scale was very difficult for the patients to understand and answer. As there is no standard translation of the DSM V scale available in Urdu or Pashto, we had to translate each question to the best of our ability, so there may have been some misinterpretation of the questions by the patients. Some of the patients, especially the females, were reluctant in responding to the questions they were asked, and might have given biased answers which could have masked the reality of their situation.

## CONCLUSION

### Summary

The study conducted showed that prevalence of PTSD in lower age groups was the low while those in their 30s and 40s had higher prevalence. One apparent reason might be their obligatory duties like earning bread and butter for their families, which they wouldn’t be able to do after their injury. Those who had lumber lesions had higher prevalence as well which might be due to social problems like uncontrolled defecation and urination. The study also demonstrated that those falling from height had higher prevalence of PTSD; the reason could be the severity of trauma sustained while falling from height, as well as the horrifying experience of free-falling.

### Recommendations

The Paraplegic Center in Hayatabad Peshawar is doing an outstanding job for these patients; more centers like these are needed throughout KPK, as there are a lot of patients who suffer from traumatic spinal cord injuries and there is no facility to treat them. These facilities should be equipped with full-fledged in-house psychiatric services, to minimize the mental effects of the patients’ injuries. Vocational training can be provided to these patients, as it is done at the Paraplegic Center in Peshawar, to keep the patients occupied and equip them with a skill that they can use to make a living. Even if the majority of these patients are not diagnosed with PTSD, the greater majority still has a number of mental health issues such as anxiety and depression due to their injuries. There is no question that mental health rehabilitation for these patients is just as important as their physical rehabilitation. The Paraplegic Center is a pioneer in this regard in the whole province, and more centers like this need to be established to serve the people who are in dire need of them.

## Data Availability

All data produced in the present work are contained in the manuscript.

## LIST OF ABBREVIATIONS

PTSD Post Traumatic Stress Disorder
WHO World Health Organization
DSM-5 Diagnostic and Statistical Manual of Mental Disorders Fifth Edition
KPK Khyber Pakhtunkhwa
DSM-III-R Diagnostic and Statistical Manual of Mental Disorders Third Edition Revised

## Appendix A Post-Traumatic Stress Disorder in Patients with Traumatic Spinal Cord Injuries

### Questionnaire (DSM-5)

Name: Date of injury:

Age: Cause of injury:

Sex: Level of injury:

Region: Date of admission:

### Criterion A: stressor

The person was exposed to: death, threatened death, actual or threatened serious injury, or actual or threatened sexual violence, as follows: **(one required)**

1. Direct exposure.
2. Witnessing, in person.
3. Indirectly, by learning that a close relative or close friend was exposed to trauma. If the event involved actual or threatened death, it must have been violent or accidental.
4. Repeated or extreme indirect exposure to aversive details of the event(s), usually in the course of professional duties (e.g., first responders, collecting body parts; professionals repeatedly exposed to details of child abuse). This does not include indirect non-professional exposure through electronic media, television, movies, or pictures.

### Criterion B: intrusion symptoms

The traumatic event is persistently re-experienced in the following way(s): **(one required)**

1. Recurrent, involuntary, and intrusive memories. Note: Children older than six may express this symptom in repetitive play.
2. Traumatic nightmares. Note: Children may have frightening dreams without content related to the trauma(s).
3. Dissociative reactions (e.g., flashbacks) which may occur on a continuum from brief episodes to complete loss of consciousness. Note: Children may reenact the event in play.
4. Intense or prolonged distress after exposure to traumatic reminders.
5. Marked physiologic reactivity after exposure to trauma-related stimuli.

### Criterion C: avoidance

Persistent effortful avoidance of distressing trauma-related stimuli after the event: **(one required)**

1. Trauma-related thoughts or feelings.
2. Trauma-related external reminders (e.g., people, places, conversations, activities, objects, or situations).

### Criterion D: negative alterations in cognitions and mood

Negative alterations in cognitions and mood that began or worsened after the traumatic event: **(two required)**

1. Inability to recall key features of the traumatic event (usually dissociative amnesia; not due to head injury, alcohol, or drugs).
2. Persistent (and often distorted) negative beliefs and expectations about oneself or the world (e.g., “I am bad,” “The world is completely dangerous”).
3. Persistent distorted blame of self or others for causing the traumatic event or for resulting consequences.
4. Persistent negative trauma-related emotions (e.g., fear, horror, anger, guilt, or shame).
5. Markedly diminished interest in (pre-traumatic) significant activities.
6. Feeling alienated from others (e.g., detachment or estrangement).
7. Constricted affect: persistent inability to experience positive emotions.

### Criterion E: alterations in arousal and reactivity

Trauma-related alterations in arousal and reactivity that began or worsened after the traumatic event: **(two required)**

1. Irritable or aggressive behavior
2. Self-destructive or reckless behavior
3. Hypervigilance
4. Exaggerated startle response
5. Problems in concentration
6. Sleep disturbance

### Criterion F: duration

Persistence of symptoms (in Criteria B, C, D, and E) for more than one month.

### Criterion G: functional significance

Significant symptom-related distress or functional impairment (e.g., social, occupational).

### Criterion H: exclusion

Disturbance is not due to medication, substance use, or other illness.

